# Modeling Strategic Interactions in Recreational Cannabis Legalization: An Evolutionary Game Approach to Market Dynamics

**DOI:** 10.1101/2025.09.15.25335743

**Authors:** Mu Wang, Jingjing Ding, Zhenkun Liu, Guangwei Deng, Yanyan Chen

**Author notes:** Corresponding Author, Address: Hubin Building, Science Island, Shushan, Hefei, Anhui, 230000,PR China. Author contributions: Mu Wang: Conceptualization, Funding acquisition, Investigation, Methodology, Writing – original draft Zhenkun Liu and Yanyan Chen: Investigation, Validation, Formal analysis Guangwei Deng and Jingjing Ding: Writing – review and editing. Declarations of competing interest: The authors declare no competing interests.

## Abstract

**Background and Aims:** The global shift toward cannabis legalization raises urgent policy challenges regarding the transition from illicit to regulated markets. This study constructs an evolutionary game-theoretic model to investigate the strategic interactions between local governments and cannabis dealers under dynamic market conditions.

**Methods:** The model assumes two players: the government, which chooses between prohibition and legalization, and dealers, who choose between legal or illicit trading. Payoffs are determined by enforcement costs, tax revenues, public health expenditures, and market scale. The growth of legal and illegal markets is modeled using logistic and exponential functions, respectively.

**Findings:** Simulations reveal convergence to a low-intensity mixed-strategy equilibrium where government reduces enforcement and dealers partially shift strategies. Under this equilibrium, minimal enforcement persists alongside a small but stable illegal market share. Sensitivity analysis shows that the decay rate of the illegal market and the expansion rate of the legal market significantly affect the speed of convergence, whereas tax rate changes exert limited influence.

**Conclusions:** The findings suggest that expanding legal market access and enhancing regulatory infrastructure are more effective than relying solely on punitive measures or tax increases. The proposed model offers policymakers a flexible tool to anticipate market responses under various cannabis legalization strategies. Beyond cannabis, the proposed framework can be generalized to other highly regulated industries undergoing formalization, such as alcohol, tobacco, or emerging digital economies.

## 1. Introduction

The global debate over cannabis legalization extends across legal, economic, and public health dimensions. Countries such as Canada, Uruguay, and parts of the United States have embraced legalization, aiming to regulate the market and obtain potential benefits. In contrast, others including China, Indonesia, and Singapore, maintain strict prohibitions, treating cannabis-related activities as serious criminal offenses[1]. This divergence in policy reflects different national priorities. For example, some governments emphasize public health and law enforcement, while others focus on economic opportunities and civil liberties.

Opinions on cannabis legalization remain deeply divided. Advocates argue that the benefits of cannabis legalization outweigh the drawbacks, while others hold the opposite view. Proponents of legalization argue that regulated markets can generate substantial tax revenue, and job opportunities[2]. For instance, states in the U.S. that have legalized cannabis have seen the increase of tax income, house prices and population[3]. Additionally, legalization may reduce the burden on the judicial system by lowering the violent crime[4]. For example, the cannabis related arrests have significantly decreased[5]. On the other hand, opponents warn that cannabis use could lead to higher cannabis use related disorders, such as psychiatric issues and addiction[5]. Public health concerns remain at the forefront of the discussion, particularly regarding the potential for cannabis to serve as a gateway drug for harder substances[6]. Moreover, some even claim that the tax revenue increased in the cannabis market is the result of the decrease of other market, such as alcohol and cigarette[7]. These contrasting viewpoints illustrate the complexity of the issue and the challenge of finding a balanced policy approach.

The strategic interaction between governments and cannabis market participants can be effectively modeled through indirect evolutionary game theory, which captures how behavioral preferences and strategies evolve under changing payoffs[8]. Unlike classical game theory, which assumes perfectly rational agents and static incentives, EGT incorporates bounded rationality, adaptive learning, and endogenous feedback loops. This makes it particularly suitable for analyzing institutional transitions in highly regulated markets, where actors adjust not only to policy rules but also to the evolving strategies of other actors under uncertainty. EGT has been widely applied in policy domains such as renewable energy subsidies[9] and environmental regulation[10], demonstrating its usefulness in capturing the dynamic interplay between enforcement, compliance, and market adaptation.

One of the central challenges in the legalization of previously prohibited markets is managing the transition from informal to formal economic structures. The persistence of an illicit market may undermine regulatory objectives by sustaining parallel supply channels and distorting competition. In this context, our evolutionary game model provides a tool to anticipate not only the pace of market convergence but also the broader policy trade-offs between enforcement, taxation, and infrastructure investment. Beyond cannabis, the framework highlights generalizable mechanisms of compliance that are relevant for other highly regulated sectors undergoing institutional transformation.

This study aims to explore following questions:

1. Can evolutionarily stable strategies (ESS) provide predictive insights for both policymaker and cannabis dealers in legal or illegal markets?
2. How do tax policies, legal and illegal markets size and other parameters shape long-term market equilibrium?
3. What strategic choices are optimal for local governments and cannabis dealers after legalization?
4. What temporal dynamics influence optimal entry windows for legal market participants?

## 2. Methods-Construction of evolutionary game model

### 2.1.1 Assumptions of evolutionary game model

This section outlines the mathematical structure of the evolutionary game model, including the key parameters, payoff functions, and market evolution dynamics. To construct an evolutionary game model that captures the dynamic interactions between government regulators and illegal cannabis market participants, we make the following assumptions based on both empirical reality and evolutionary modeling practices:

1. The model involves two primary players: In one province or state, for now, there are illegal dealers of cannabis. The government determines whether to support cannabis legalization or maintain strict prohibition. Its strategic behavior is expressed by the probability x∈ [0,1], where x=1 indicates 100% prohibition (strict enforcement) and x=0 full legalization, indicating the preference for legalization. Cannabis dealers, representing existing unlicensed dealers who choose either to remain in the black market or enter the legal market, and potential dealers who have not decided whether to enter the market. Their strategy is denoted by probability y∈ [0,1], where y=1 indicates 100% illegal operations and y=0 full legal compliance, reflecting the propensity to remain in the illegal market. Government-run cannabis stores are not considered in this model. Their decisions and payoffs are shown in Figure 1.
2. The model includes following cost: The cost of operating in the legal market includes licensing fees, regulatory compliance, and competitive pressure, which is assumed to be non-trivial. The cost of operating in the illegal market includes the risk of law enforcement and financial penalties. Legal cannabis sales generate taxation revenues for the government, proportional to a tax rate S, while illegal activity generates no fiscal benefit and adds enforcement costs.
3. Dynamic, time-dependent variables: Key variables, including law enforcement costs, market demand, and public health expenditures, are modeled as time-dependent, influenced by the evolving strategies of governments and dealers. The simulation spans 24 months, starting from Month 1.
4. Adaptive strategy evolution and incomplete rationality The players do not make one-shot, fully rational decisions. Instead, their strategies evolve over time based on historical payoffs, using replicator dynamics. This aligns with indirect evolutionary game theory, where behavioral rules are shaped by success and environmental feedback rather than perfect foresight.
5. Existence of evolutionary stable strategy (ESS) Under suitable parameter conditions, the system is assumed to converge toward one or more ESS, where neither player benefits from unilaterally deviating. These equilibria represent long-run policy-market outcomes.

**Figure 1.**
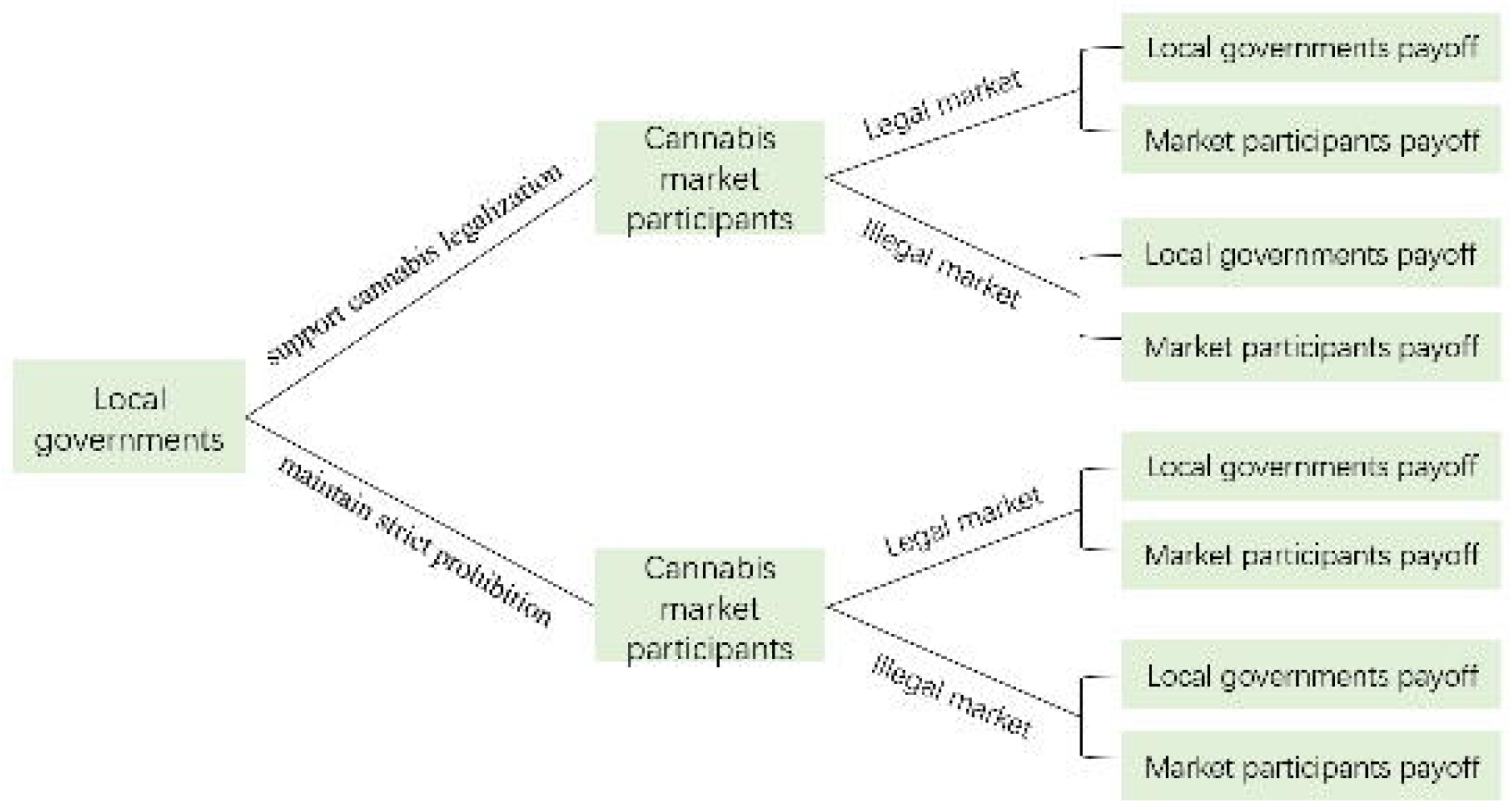
demonstration of decisions and payoffs for both actors

### 2.1.2 Parameters

In this study, parameter values were selected based on real-world data from jurisdictions that have implemented cannabis legalization, primarily drawing from post-legalization experiences in Canada and selected U.S. states. The values are composite estimates and not derived from a single unified dataset. Therefore, the model serves as a stylized representation of strategic dynamics, rather than a direct simulation of any specific jurisdiction. In addition, in this study, the scenario is set in a state or province where cannabis legalization has not yet been implemented, and all parameter settings are measured on a monthly basis.

The initial price for cannabis in legal stores and illegal stores is $13.08 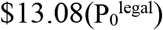 and $10.23(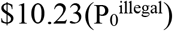) per gram, respectively, based on the real-world data from two months post-legalization in Canadian market [11]. The cost of cannabis set $2 per gram for legal stores, based on the real world data[12] and 1.5 per gram for illegal dealers. Given the licensed wholesalers and retailers had reached 1.4 billion grams total in Canada[12], we set the initial market size 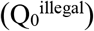 was 1 million (grams/units) per month pre-legalization and it was illegal. Since the cannabis use would increase in adults after legalization[13], the total market size (Q_max_) increased to 1.3 million (grams/units) per month and meanwhile, the legal market will increase and gradually take over the illegal market[14]. The illegal market would gradually decrease, because of the growth of the legal market[15], but reserves a small portion of the market. Given the illegal market still exists, even if the cannabis trade is legal[16], we set the minimum illegal market (Q_min_) to 320000 (grams/units), around 25% of the total market, based on the real world data in Canada[17]. The tax rates (S) were initially set to 0.10, but increased to 0.14 and to 0.18 in parameters sensitivity test. According to the real tax rates in New Mexico, in the first three years after legalization the tax rate was 0.12 and increased one percentage point annually until it reached 0.18[18]. We also chose a higher tax rate (0.3) to test whether high tax rate would interfere with the growth of the legal market. The initial cost of law enforcement (C_f_) was $4500, based on the fiscal data from Canada’s government for fighting cannabis related crime over five years in 2017[19]. The public health issues caused by more use of cannabis, such cannabis related hospitalization, has been warned by researchers[5] are assigned a baseline value of was set $100000 (H_0_). Financial support (I) from national programs, such as the Byrne Justice Assistance Grant, is modeled as $50000. Other detailed parameter settings are shown in Table 1.

**Table 1.** Parameters in evolutionary game model

| Parameters | Symbol | Initial Value | Unit | Parameter Description |
| --- | --- | --- | --- | --- |
| Government's Choice Probability | $x$ | - | - | Range [0,1], where 1 indicates a 100% probability of prohibiting cannabis legalization. |
| Cannabis Dealers' Choice Probability | $y$ | - | - | Range [0,1], where 1 indicates a 100% probability of engaging in illegal operations. |
| Initial Government Enforcement Cost | $C_0$ | 4500 | USD | Initial cost of government enforcement. |
| Enforcement Cost Adjustment Rate | $\delta$ | 10 | Dimensionless | Rate of change in enforcement costs. |
| Financial assistance for anti-drug efforts | $I$ | 50000 | USD | Local governments can receive financial assistance from national or international bodies. |
| Illegal Market Initial size | $Q_0^{\text{illegal}}$ | 1 | Million units | Initial sales volume in the black market. |
| Illegal Market Minimum Sales | $Q_{\min}$ | 0.32 | Million units | Minimum sales volume in the black market. |
| Illegal Market Decay Rate | $\alpha$ | 0.1 | Dimensionless | Decay rate of the illegal market due to enforcement and competition. |
| Total market size post-legalization | $Q_{\max}$ | 1.3 | Million units | Maximum sales capacity of the legal market. |
| Legal Market Opening Time | $T_0$ | 1 | Month | Time when the legal market is introduced (year). |
| Legal Market Growth Rate | $\beta$ | 0.5 | - | Expansion speed of the legal market. |
| Legal Market Price | $P_0^{\text{legal}}$ | 13.08 | USD/gram | Price of cannabis in the legal market and this price was reduced the cost. |
| Illegal Market Price | $P_0^{\text{illegal}}$ | 10.23 | USD/gram | Price of cannabis in the illegal market and this price was reduced the cost. |
| Government Tax Rate | $S$ | 10 | Percentage | Tax rate imposed on the legal market. |
| Additional Public Health Cost | $H_0$ | 100000 | USD | Initial public health expenditure. |
| Health Cost Growth Coefficient | $\eta$ | 0.005 | - | Coefficient of health cost growth impact on market size. |
| Enforcement Cost | $C_f$ | - | - | The initial cost of law enforcement was \$4500, but changing based on illegal market size. |
| Legal Market Entry Cost | $v$ | 2000 | USD | Average monthly cost for licenses and compliance in the legal market. |
| Legal Cost per Gram | - | 2 | USD/gram | Production cost per gram in legal market |
| Illegal Cost per Gram | - | 1.5 | USD/gram | Production cost per gram in illegal market |
| Illegal Operation Cost | $F$ | 1 | USD | Risk cost per gram of cannabis sold in the illegal market. |

### 2.1.3 Assumption of parameters that may affect both agents

In this study, we assume that the sales volume and price of cannabis in the legal and illegal markets are different and dynamically changing. Given our prior assumption that the local government has not yet implemented cannabis legalization, the sales volume in the legal market is set to zero at the initial time point (time = 0). But the demand will increase as legal market growing. The logistic growth model for Q captures the S-shaped diffusion curve emphasized in new product marketing research[20], reflecting the constrained expansion of legal cannabis markets post-legalization. The equation is:

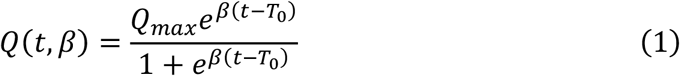

Where *Q*_*max*_ is the maximum legal market size; β represents the legal market expansion rate. *T*_0_=1 represents the time when the legal market is introduced.

The illegal market demand follows an exponential decay model, as increasing law enforcement and legal alternatives reduce its size:

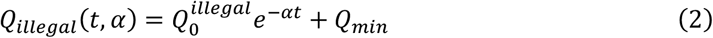

Where *Q*_*min*_ is the minimum illegal market of Cannabis, because the illegal market will still exist, even if the cannabis trade is legal; 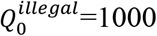 is the initial illegal market size. α is the illegal market decay rate; influenced by enforcement and economic factors.

When combating criminal activities, the government incurs law enforcement cost, referring ‘*C*_*f*_‘, including increased police deployment, equipment procurement, and other related expenses. *C*_*f*_ decreases as the illegal market shrinks, given by:

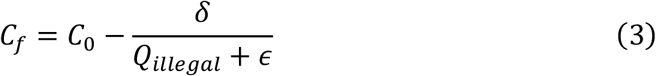

Where *C*_0_ is the initial enforcement cost, and δ controls the enforcement cost adjustment rate.

These payoff structures are first-order approximations and facilitate the tractable use of replicator dynamics in strategy evolution modeling.

#### 2.2 Local government and drug dealers’ strategy choices

This study models a two-player evolutionary game between the government and drug dealers, where each player continuously adjusts their strategy based on payoffs as time goes. For local governments, they can choose to fight the illegal cannabis market through a strict enforcement (SE) strategy, in which all cannabis transactions are considered illegal. Alternatively, they can choose a cannabis-legalized (CL) policy, allowing cannabis trade to become a lawful economic activity within the region, while the illegal market may still persist. Dealers can engage in illegal trade (IT) if cannabis remains prohibited, or choose between legal trade (LT) and continued illegal activity under legalization.

### 2.2.1 Expected Payoff Functions

Based on local governments’ strategy, the payoff of them can be from strategy SE (*U*_*SE*_) or CL(*U*_*CL*_) and their payoff equations are:

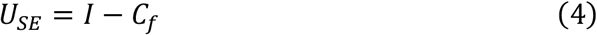

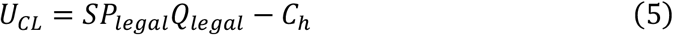

Thus, based on equation (4) and (5), their expected utility is given by:

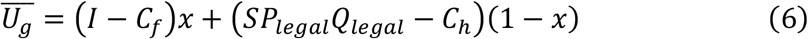

Based on drug dealers’ strategy, the payoff to them can be from strategy IT (*U*_*IT*_) or LT (*U*_*LT*_) and their payoff equations are:

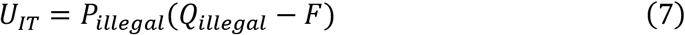

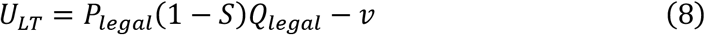

Thus, based on equation (7) and (8), the expected utility of drug dealers is

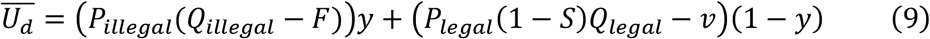

### 2.2.2 replicator dynamics

To model the dynamic evolution of strategic behaviors between the local government and cannabis dealers, we adopt the framework of replicator dynamics, a foundational tool in evolutionary game theory. This approach assumes that agents do not necessarily make fully rational decisions in the classical sense, but instead adapt their strategies over time based on relative payoff performance. Therefore, the rate of change in strategy choice is proportional to the payoff advantage over the population mean, as reflected in the replicator dynamics applied below. Based on (4)-(6), we have the corresponding replicator dynamic equation for government:

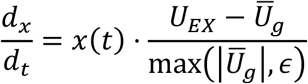

Based on (7)-(9), we have the corresponding replicator dynamic equation for cannabis dealers:

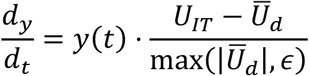

The denominator of the equation represents the average payoff across the population, making the rate of change proportional to the strategy’s payoff advantage.

#### 2.3 Parameters sensitivity analysis

To evaluate how policy design influences the strategic dynamics between regulators and cannabis dealers, we conducted a sensitivity analysis on three key parameters: the tax rate S, the decay rate of the illegal market α, and the expansion rate of the legal market β. Specifically, the tax rate S reflects the fiscal burden or incentive within the legal system and affects both the government’s revenue from the legal market and dealers’ willingness to transition into it. The decay rate α models the speed at which the illegal market erodes over time due to enforcement or loss of profitability, shaping the enforcement return profile for the government. β characterizes how fast the legal infrastructure expands, influencing how quickly legal channels become accessible and attractive to dealers. Analyzing these parameters allows us to examine not only equilibrium outcomes but also transitional behaviors.

## 3 Simulation results

Figure 2 illustrates the dynamic evolution of strategic behaviors for both the local government and cannabis dealers under varying initial conditions. Panel (a) illustrates that the government’s enforcement probability x(t) declines rapidly, eventually stabilizing around a low level. This reflects a rational withdrawal from costly enforcement as marginal returns diminish. Panel (b) reveals that dealers’ probability y(t) of remaining illegal initially plateaus or slightly increases, which may due to temporarily exploit regulatory gaps or delay switching due to high transition costs. However, as legal markets expand and illegal profits fall, this strategy becomes less viable. Over time, y(t) declines and converges to a similar low equilibrium around y≈0.05. The simulation system converges to a low-intensity mixed-strategy equilibrium, where minimal enforcement coexists with a small illicit segment, a characteristic of an ESS.

**Figure 2.**
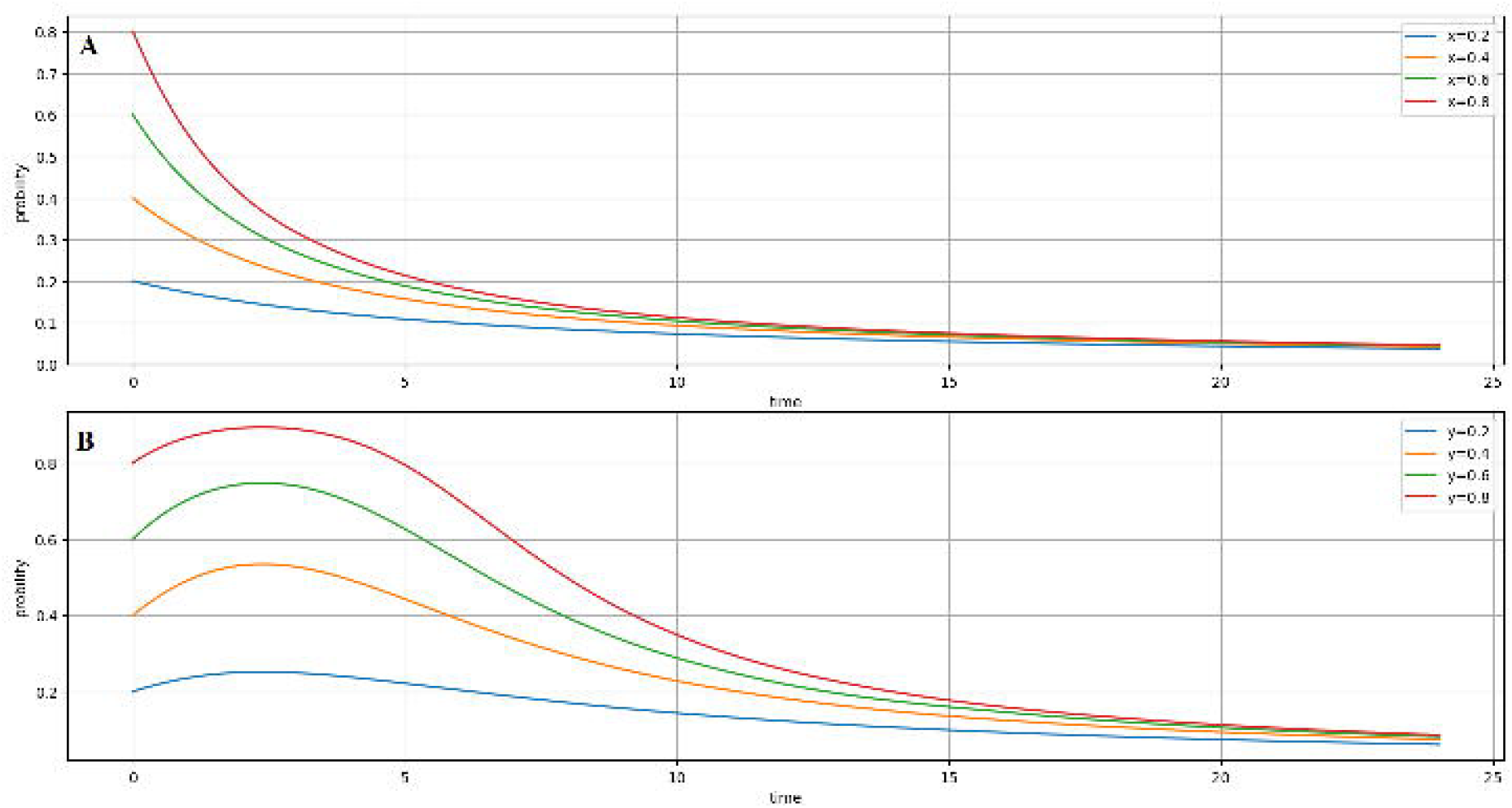
Evolutionarily Stable Strategy over time in different probability. Dynamic evolution of government enforcement strategy x(t) and dealer behavior y(t) over time under varying initial conditions. Panel (a) shows that x(t) consistently declines toward a low level, while panel (b) illustrates that y(t) also converges to a low equilibrium after minor fluctuations. Both trends indicate convergence to a low-intensity evolutionarily stable strategy (ESS).

To explore how key environmental and policy parameters influence the strategic interaction between governments and illicit cannabis dealers, we simulate the system’s dynamic responses to variations in the legal market expansion rate (β), the illegal market decay rate (α), and the cannabis tax rate (S). Figures 3–5 present both the temporal evolution of strategies over time and their phase trajectories in the x(t) and y(t) state space. The illegal market decay rate and the legal market expansion rate are key as exponential factors in market dynamics, controlling erosion and expansion speeds more than linear terms like taxes.

**Figure 3.**
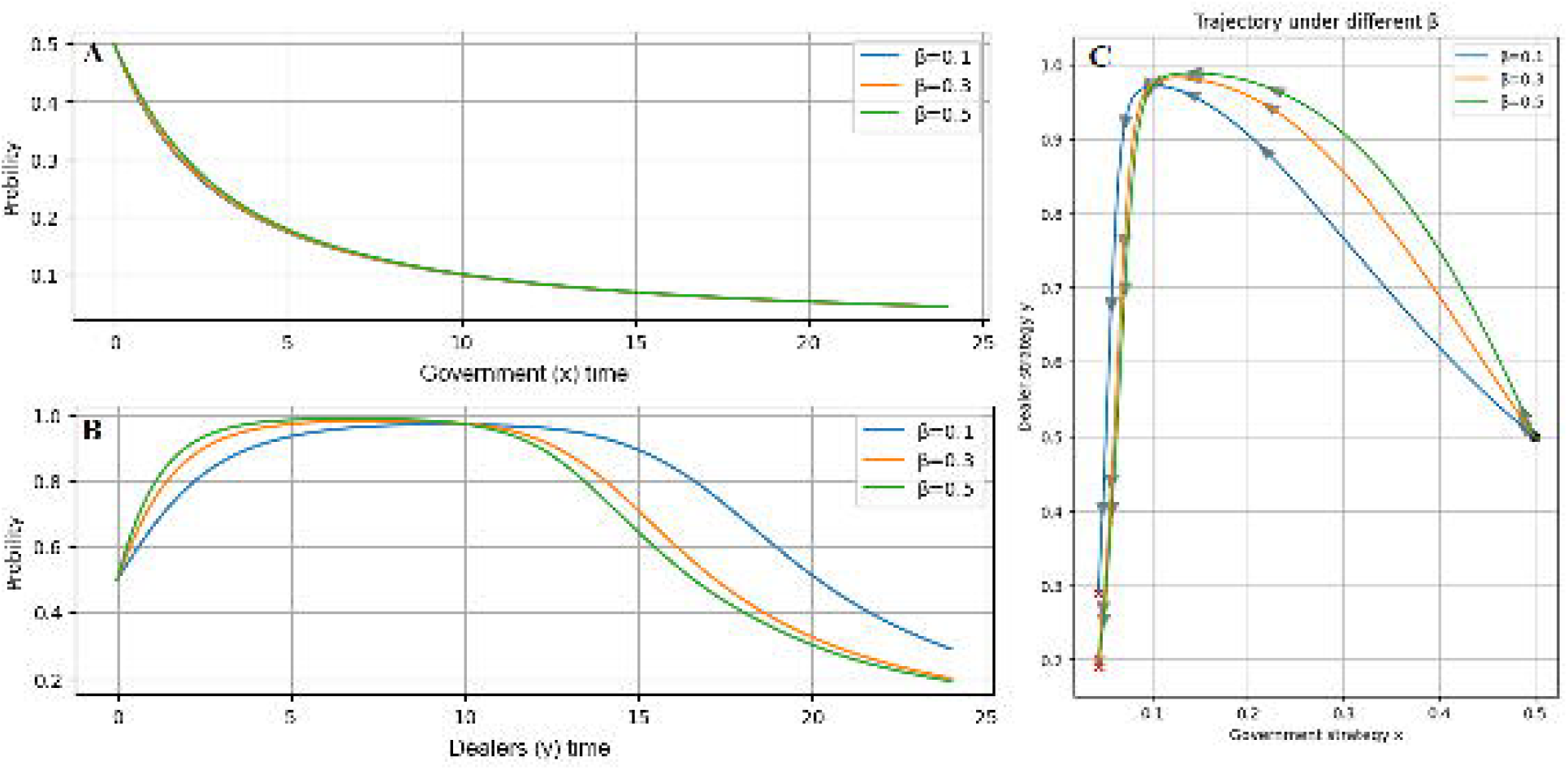
Effects of Legal Market Expansion Rate (β) on Strategic. Dynamics Sensitivity of strategy evolution to different values of β (legal market expansion rate). Faster legal market growth accelerates dealer transition to legality (y(t) decline) and enables earlier stabilization of government enforcement (x(t)). Phase plots demonstrate steeper convergence paths under higher β.

The results reveal that the model is more sensitive to structural parameters such as α and β than to fiscal instruments like S. In Figure 3, which examines the influence of β, we observe that higher values of β, representing faster expansion of the legal cannabis market, lead to a steeper and earlier decline in the probability that dealers continue operating in the illicit sector y(t). While the government’s enforcement probability x(t) remains relatively stable across β values, the trajectory plots (panel C) suggest that faster legal market growth enables earlier stabilization at lower enforcement intensities. These results highlight the role of legal infrastructure development in crowding out illegal activity.

Figure 4 illustrates the effects of α, the decay rate of the illegal market. Higher α values induce steeper and faster decline in y(t), indicating that rapid contraction of illicit market volume accelerates dealer withdrawal from illegal trade. The government’s response is also slightly more active at early stages under higher α, as seen in both time series and trajectory plots, but enforcement effort tapers off more quickly once market signals stabilize. These dynamics underscore the strategic interdependence between enforcement credibility and market erosion speed: when the illicit market naturally declines, even modest policy intervention can guide the system toward compliance.

**Figure 4.**
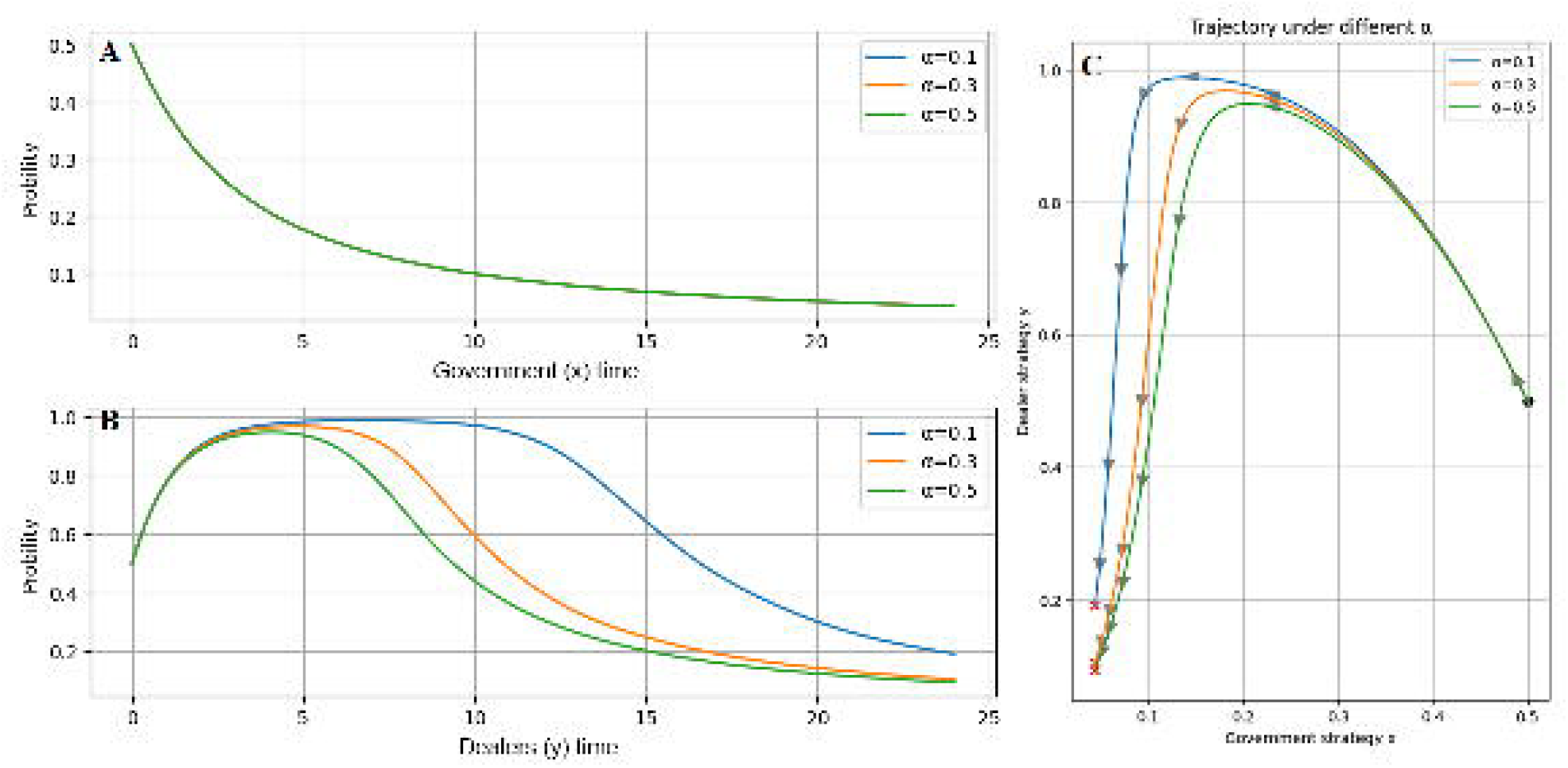
Effects of Illegal Market Decay Rate (α) on Strategic. Dynamics Sensitivity of strategic dynamics to changes in α (illegal market decay rate). A higher α leads to quicker withdrawal of dealers from illicit trade and slightly more proactive government enforcement early in the process. Trajectories converge faster under stronger illegal market decline.

By contrast, Figure 5 shows that changes in the cannabis tax rate S exerts only marginal influence on both x(t) and y(t). Across all tax scenarios, strategy paths converge to similar equilibrium values, and only minor shifts in path curvature are observed. This confirms that taxation may influence fiscal revenues but has minimal impact on strategic behaviors, highlighting the limited role of tax rate alone in displacing illicit trade. This reflects high tax burdens alone cannot displace illegal markets without strong legal competitiveness and enforcement.

**Figure 5.**
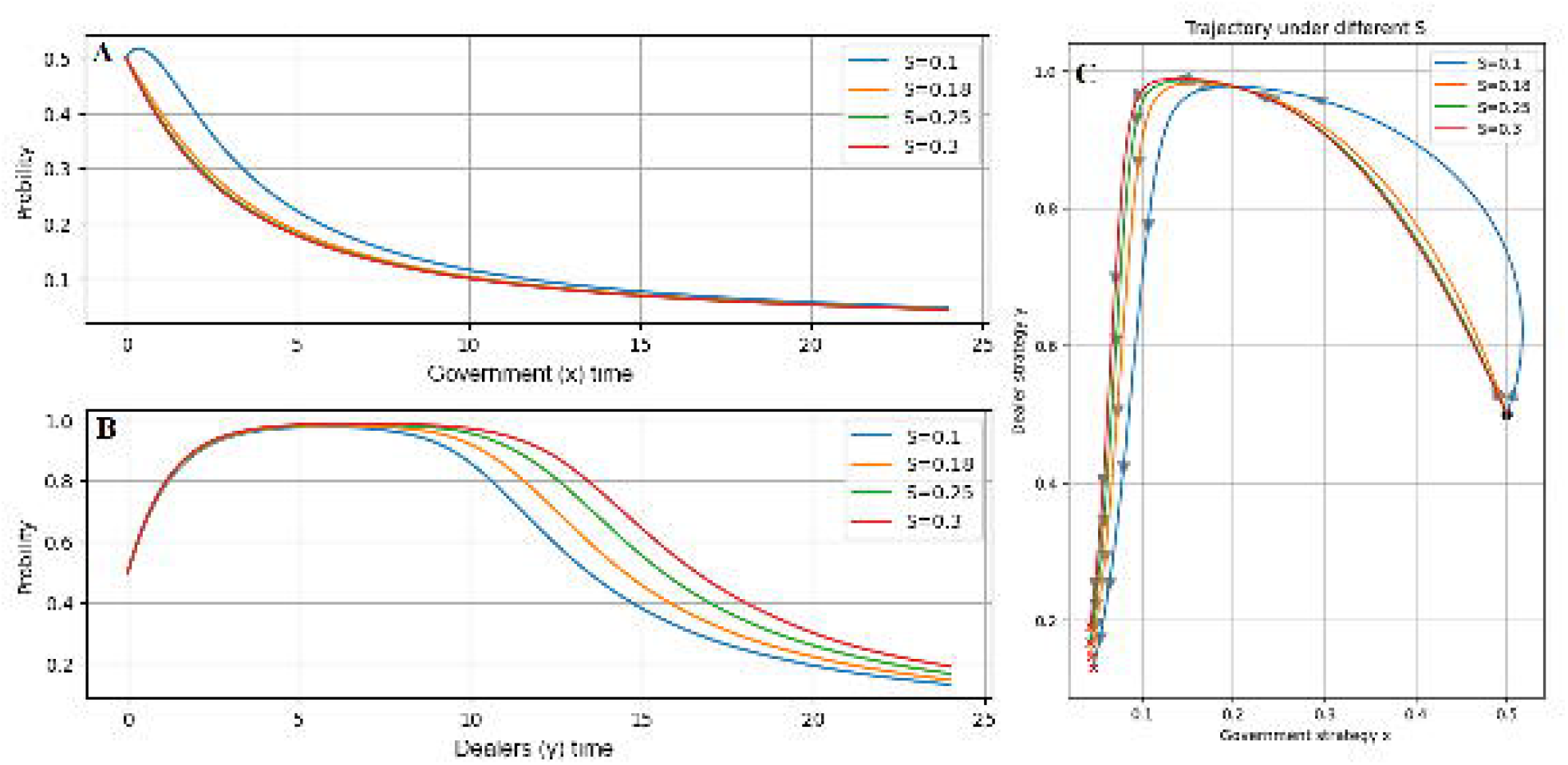
Effects of Cannabis Tax Rate (S) on Strategic Dynamics. Effects of varying tax rates (S) on strategy trajectories. Despite changes in taxation, both x(t) and y(t) follow similar paths and converge to nearly identical equilibria. This suggests that taxation alone has limited influence on the strategic behavior of governments and dealers.

## 4 Discussion

This study employs evolutionary game theory to examine the strategic interactions between government regulators and cannabis market participants. Through dynamic simulations and parameter sensitivity analysis, the research reveals not only the stability of long-term equilibria but also the behavioral mechanisms that drive convergence. The results suggest that tax policies show limited impact on long-term strategic behavior. Instead, market structure dynamics, particularly the decay rate of the illicit sector and the growth rate of the legal sector, are more influential. In essence, the pace of market transformation or the relative scale of the legal market outweigh taxation in shaping outcomes. Rapid expansion of legal supply channels, coupled with the natural erosion of black-market infrastructure, facilitates smoother and more effective transitions.

For policymakers, prioritizing the expanding and protection of the legal market should take precedence over punitive enforcement is essential. Adaptive strategies, such as early investment in legal infrastructure and selective enforcement against illegal actors, such as targeting high-risk dealers rather than widespread punitive actions, are more effective than punishment. Governments can lower initial entry barriers by reducing license fees or simplifying procedures to attract more participants into the legal sector, thereby accelerating its growth and compressing the space for illicit competition. New Mexico has implemented a similar policy in taxation. Their tax was settled at 12 percent in the first three years and increased by one percentage point annually until it reached 18 percent [18]. This strategy created breathing space for legal businesses to grow before facing full fiscal pressure. Using tax revenue to support compliance enforcement, rather than over-penalization, may also yield more sustainable outcomes. In contrast, complex regulatory processes, as an example of California’s illegal market, may inhibit the growth of legal market and perpetuate illegal market[21].

Institutional theory emphasizes that policy adoption depends not only on legal frameworks, but also on legitimacy, normative alignment, and cognitive acceptance [22]. Early expansion of the legal cannabis market can generate a legitimizing effect, building consumer trust and normalizing compliance. As legal infrastructure grows, behavioral inertia and market feedback loops begin to reinforce lawful participation, reducing the need for intensive enforcement. This reflects a path-dependent transition [23], where initial investments in institutional support shape long-term actor behavior. Once the legal sector achieves sufficient scale and credibility, tax incentives become less critical, as compliance is sustained by structural alignment. As legal infrastructure grows, it also fosters healthier consumer behaviors by reducing access to adulterated illicit products, potentially lowering addiction prevalence and related healthcare costs. These dynamics support a shift from enforcement-led to infrastructure-led legalization, in which governments actively shape markets through early support rather than coercion.

Based on these findings and Scott’s (2013) three pillars of institutions, regulative, normative, and cognitive[22], we argue that enforcement mechanisms primarily activate the regulative pillar, while infrastructure investment operates on the normative and cognitive dimensions. This distinction helps explain why infrastructure-led strategies demonstrate stronger long-term effectiveness. To better capture this process in the context of cannabis legalization, we develop a Three-Phase Legalization Compliance Model: 1) Phase 1 (prohibition breakdown), targeted enforcement destabilizes the illicit market and initiates behavioral disruption. 2) Phase 2 (market channeling) occurs as legal infrastructure expands, reducing compliance costs and building perceived legitimacy, which begins to attract participants toward lawful behavior. 3) Phase 3 (compliance consolidation), in which behavioral inertia and institutional credibility reinforce lawful strategies, making deviation socially and economically irrational. These phases reflect a dynamic path-dependent process, in which early structural choices shape long-term behavioral equilibria. As such, our results extend institutional path dependence theory from Sydow et al. (2009) by incorporating explicit actor strategy adaptation in a market-based context[23].

Moreover, building on the simulation’s revelation of initial fluctuations in dealer strategies, an additional insight arises from asymmetric risk preferences: risk-averse actors may delay transitions because perceived enforcement losses outweigh potential gains from legal entry[24]. This behavioral mechanism, consistent with prospect theory, explains early plateaus or temporary increases in illicit activity. It highlights how bounded rationality amplifies market differentiation initially[25], underscoring the need for policies that reduce perceived transition costs to accelerate convergence.

For legal cannabis operators, market credibility, ease of entry, and price competitiveness are critical. Simulation trajectories show that when legal profitability outweighs the cost of compliance, dealers gradually transition away from the illicit market as compliance becomes economically viable. Policies that facilitate licensing, reduce startup barriers, and stabilize pricing are instrumental in anchoring this shift. Policies that facilitate licensing, reduce startup barriers, and stabilize pricing are instrumental in anchoring this shift. This transition also enhances public health by ensuring regulated, high-quality cannabis, reducing risks of contamination-related illnesses that affect vulnerable populations. In addition, simulation results suggest that the optimal entry point for legal cannabis dealers lies in the early-to-middle phase of legal market expansion, when illicit competition begins to wane but regulatory support and market space remain favorable. Entering too early exposes operators to risk from unregulated competition, while entering too late may lose market share and institutional advantages.

## Data Availability

It is a health policy realted artical. Authors used model simulation, without dataset.

## Limitations

This study, while offering a dynamic framework to analyze strategic interactions in cannabis markets, has several limitations. First, the model assumes homogeneous agents within each group, but ignores the intra-group heterogeneity such as regional enforcement variation or diverse dealer risk preferences. Second, government-run stores of cannabis and consumers were not discussed in this article. More agents could be involved in a Tripartite Evolutionary Game in future study.

This study focuses exclusively on recreational cannabis legalization policies, excluding medical cannabis, which involves distinct regulatory frameworks and is beyond the current scope. We aim to explore the strategic interactions between local governments and cannabis dealers using an evolutionary game approach.

## Declaration of generative AI in scientific writing

During the preparation of this work the authors used Open AI ChatGPT in order to polish the written English and improve its readability only. After using this tool, the authors reviewed and edited the content as needed and take full responsibility for the content of the published article.

## Notes

Fundings: This work is supported by National Natural Science Foundation of China, grant 72188101

### Competing Interest Statement

The authors have declared no competing interest.

### Funding Statement

This work is supported by National Natural Science Foundation of China, grant 72188101.

## References

[1] Global Landscape of Marijuana Legislation. 2024

[2] Gallien M, Occhiali G. Cannabis taxation as a revenue source for development: Opportunities and challenges. 2023

[3] Brown J, Cohen E, Felix RA. Economic benefits and social costs of legalizing recreational marijuana. Federal Reserve Bank of Kansas City Working Paper. 2023:23–10

[4] Dills AK, Goffard S, Miron J, Partin E. The effect of state marijuana legalizations: 2021 update. Cato Institute, Policy Analysis. 2021

[5] Hall W, Stjepanović D, Dawson D, Leung J. The implementation and public health impacts of cannabis legalization in Canada: a systematic review. Addiction. 2023;118:2062–72

[6] Wilson J, Mills K, Freeman TP, Sunderland M, Visontay R, Marel C. Weeding out the truth: a systematic review and meta-analysis on the transition from cannabis use to opioid use and opioid use disorders, abuse or dependence. Addiction. 2022;117:284–98

[7] Miller K, Seo B. The effect of cannabis legalization on substance demand and tax revenues. National Tax Journal. 2021;74:107–45

[8] Sandholm WH. Evolutionary game theory. Complex social and behavioral systems: game theory and agent-based models. 2020:573–608

[9] Yang Y, Yu X, Wang B. Evolutionary Game Analysis of Multi-Agent Synergistic Incentives Driving Green Energy Market Expansion. Sustainability. 2025;17:7002

[10] Zhang Z, Gao Z. Can environmental regulation synergy inhibit the differentiation of the Porter effect in carbon markets? Economic Analysis and Policy. 2025

[11] Mahamad S, Wadsworth E, Rynard V, Goodman S, Hammond D. Availability, retail price and potency of legal and illegal cannabis in Canada after recreational cannabis legalisation. Drug and alcohol review. 2020;39:337–46

[12] Lamers M. Canadian wholesale cannabis prices are off more than 40% in 2022. 2023

[13] Gibbs B RT, Wride S. Cannabis Legalization—Canada’s experience. 2021

[14] Hathaway AD, Cullen G, Walters D. How well is cannabis legalization curtailing the illegal market? A multi-wave analysis of Canada’s national cannabis survey. Journal of Canadian Studies. 2021;55:307–36

[15] Amlung M, MacKillop J. Availability of legalized cannabis reduces demand for illegal cannabis among Canadian cannabis users: Evidence from a behavioural economic substitution paradigm. Canadian Journal of Public Health. 2019;110:216–21

[16] Boyle EB; Hurd YL TS. Cannabis Policy Impacts Public Health and Health Equity. Washington (DC): National Academies Press (US); 2024

[17] McDonald AJ, Cooper A, Doggett A, Halladay J, Belisario K, MacKillop J. Association of recreational cannabis legalization with changes in medical, illegal, and total cannabis expenditures in Canada. International Journal of Drug Policy. 2025;139:104793

[18] Cannabis Excise Tax. 2022

[19] Cannabis Black Market. 2020

[20] Mahajan V, Muller E, Bass FM. New product diffusion models in marketing: A review and directions for research. Journal of marketing. 1990;54:1–26

[21] Fataar F, Goodman S, Wadsworth E, Hammond D. Consumer perceptions of ‘legal’and ‘illegal’cannabis in US states with legal cannabis sales. Addictive behaviors. 2021;112:106563

[22] Scott WR. Institutions and organizations: Ideas, interests, and identities: Sage publications; 2013.

[23] Sydow J, Schreyögg G, Koch J. Organizational path dependence: Opening the black box. Academy of management review. 2009;34:689–709

[24] Zhang R, Brennan TJ, Lo AW. The origin of risk aversion. Proceedings of the National Academy of Sciences. 2014;111:17777–82

[25] Farhi E, Werning I. Monetary policy, bounded rationality, and incomplete markets. American Economic Review. 2019;109:3887–928

